# The Red Thread: Stakeholder Perspectives on Menstrual Health and Hygiene in Ghana

**DOI:** 10.1101/2025.01.22.25320983

**Authors:** Sitsofe Gbogbo, Israel Wuresah, Priscilla Klutse, Sarah Odi Mantey, Ishmael Boateng, Paramount Eli Nelson, Veronica Charles-Unadike, Darline ElReda, Constance Currier

**Affiliations:** University of Health and Allied Sciences, Ho, Ghana; Michigan State University, MI, USA

**Keywords:** menstrual health, hygiene, adolescents, Ghana

## Abstract

**Background:** Menstrual hygiene management is a challenge for girls and women in Ghana due to a lack of access to affordable menstrual products, water, sanitation, and hygiene (WASH) facilities, inadequate menstrual health education; and social stigma surrounding menstruation. This study aimed to explore the experiences and perspectives of various stakeholders and identify strategies for improving menstrual health and hygiene (MHH) among adolescent girls.

**Methods:** We recruited stakeholders for focus group discussions (FGDs) from basic schools (primary and junior high, grades 1-9) in Hohoe, Volta Region, Ghana. We conducted FGDs with adolescent boys (n=60), parents (n=48), and gatekeepers (n=19). Semi-structured guides were developed and used during FGDs, recorded and supplemented by field notes. Transcripts were thematically analyzed using MAXQDA 2024 software (1).

**Results:** The themes identified were menstrual health management, perceptions and understanding of menstruation, social and cultural practices, and education and awareness. The following strategies were identified: 1) the need for adequate wash facilities, 2) the need to address stigma associated with MHH, 3) the need to reduce the cost of sanitary materials, and 4) the need for improved menstrual health education.

**Conclusion:** These results confirm that cultural stigmas, inadequate facilities, and financial constraints still exist. These act as key barriers to MHH for schoolgirls in Hohoe, Ghana. To support girls’ health, dignity, and education, recommendations include inclusive education programs, improved WASH facilities, and affordable and/or sustainable menstrual products.

**What is already known on this topic?:** Menstrual hygiene remains a significant issue in Ghana, with challenges such as a lack of affordable menstrual products, inadequate water, sanitation, and hygiene (WASH) facilities, limited menstrual health education, and pervasive stigma.

**What this study adds:** This study uniquely incorporates perspectives from multiple stakeholders, including adolescent boys, parents, and gatekeepers (teachers, school health coordinators, and school community leaders), to provide an all-inclusive understanding of MHH barriers and strategies to address them. The study reveals a shift in cultural practices, with modernization eroding some traditions but perpetuating stigma through religious and social norms, which continue to isolate menstruating girls and women.

**How this study might affect research, practice, or policy:** The findings support policy recommendations to reduce financial barriers to menstrual products, including tax elimination or subsidies, and to invest in school infrastructure upgrades. Future research and interventions can build on this study’s insights to develop context-specific, gender-sensitive, and sustainable approaches to improving MHH.

## Introduction

Menstrual hygiene management (MHM) in Ghana presents significant challenges for girls and women, primarily due to inadequate access to affordable menstrual products, insufficient water and sanitation (WASH) facilities, inadequate knowledge about menstrual hygiene, and pervasive social stigma (1, 2, 3, 4).

### Affordability of Menstrual Products

Many girls use cloth or other materials due to the cost of sanitary products and often miss school. As a result, school absenteeism is estimated at 27.5% in rural areas in Ghana (1). Wealth disparities impact access to menstrual hygiene products, with poorer households facing more significant challenges (5). In extreme circumstances, Ghanaian girls report engagement in sexual activity to acquire funds for menstrual hygiene supplies (6).

### Sanitation and Hygiene

The lack of clean and private WASH facilities remains an issue in Ghana, affecting school attendance and overall well-being (2). Inadequate WASH infrastructure in schools is associated with absenteeism among menstruating girls in low—and middle-income countries (LMICs) (1,7,8). The availability of adequate WASH facilities and private spaces for changing results in higher attendance rates among girls (9,10,11).

### Inadequate menstrual health education and stigma

Inadequate menstrual health education contributes to misconceptions and stigma about menstruation, isolating menstruating girls (12). The absence of knowledge fosters an environment where stigma thrives, as girls internalize negative societal attitudes towards menstruation, leading to feelings of shame and embarrassment (13). As a result, girls often conceal their menstruation, hindering their participation in school and social activities; impacting their overall mental wellbeing.

Cultural restrictions and lack of support in schools hinder girls’ participation and comfort during menstruation (1). A study in Ethiopia reported that discussions about menstruation in school settings are considered taboo, leading to a lack of open dialogue between students and educators (15). Similarly, in Ghana, cultural taboos influence the understanding and management of menstruation among adolescents (15). For instance, many girls are taught that menstruation is a source of shame and impurity, leading to restrictions on their activities during this time, such as prohibitions against entering the kitchen, participating in religious activities, and even sleeping in their usual beds (17,18). These cultural practices and silence exacerbate feelings of isolation among girls, as they may feel unsupported and unprepared to manage their menstruation effectively (14). Cultural norms often discourage girls from discussing menstruation, resulting in a lack of guidance and support from peers and teachers (1). The stigma surrounding menstruation is reinforced by the attitudes of peers and educators; negative attitudes towards menstruation can lead to teasing and bullying, which further alienates girls during their menstrual cycles (19). Teachers can be the primary source of information for students regarding menstrual health. However, many educators lack the necessary training and resources to provide comprehensive menstrual health education (23,24).

In addition to challenges faced in educational settings, parents also often feel uncomfortable discussing menstruation with their children – especially with boys, leading to misinformation, inadequate preparation for menstruating girls (20,21), and further stigmatization of menstruation (22). Adolescent boys also play an essential role in the menstrual health discourse, as their attitudes and knowledge can either support or hinder the menstrual experiences of their female peers. Studies have shown that boys often receive limited education regarding menstruation, leading to misconceptions and stigma that can affect their interactions with girls during this critical period (24,25). The objective of this study was to identify strategies to improve menstrual health and hygiene experiences for school-going adolescent girls based on the unique perspectives of key stakeholders from the Hohoe Municipality in Ghana.

## Methods

### Setting

Hohoe Municipal District is one of 18 districts in the Volta Region of Ghana. The Municipal District has 91 primary and junior high schools (Ghana Education Service Hohoe, 2019). The school closest to Fred N. Binka School of Public Health, University of Health and Allied Sciences (UHAS) in Hohoe, was selected as the recruitment site to accommodate the study budget.

### Study Design and Study Population

A combination of strategies was used to recruit participants. First, announcements were made during a parent-teacher association meeting to invite interested boys and parents. Additionally, the headmaster of the school directly solicited the participation of teachers and head teachers.

This qualitative study used focus groups to collect the experiences and perspectives of boys, parents, and gatekeepers (teachers, head teachers, School Health Education Program Coordinators, and the school headmaster) from a basic school in Hohoe, Ghana, followed by a thematic analysis of the transcripts. The convenience sampling method was adopted for focus group participants. A total of 11 Focus Group Discussions (FGDs) were conducted with group sizes ranging from 7-14 participants: 5 FGDs with boys (n=60), 4 FGDs with parents (n=48), and 2 FGDs with gatekeepers (n=19).

### Data collection

Study authors developed the focus group guides used in this study. The focus group guide for the boys was based on previous work by WoMena Uganda (26) but modified for the cultural context in Ghana (Table 1). Based on the research purpose and literature review, the authors drafted additional focus group guides for use with parents and gatekeepers. We conducted pilot focus groups at a different basic school and revised the focus group guide to incorporate feedback from pilot sessions. The authors of this study conducted the focus groups. Focus groups with mothers and fathers were conducted separately to prevent passive participation, given the gender dynamics in the Ghanaian context. Each discussion started with an explanation of the purpose of the study and an invitation for the participant to complete a consent form, followed by a guided FGD. FGDs were recorded while written field journals were used to validate and supplement the data. FGDs were conducted in the English language, with some FGDs utilizing a blend of English, Ewe, and Twi to accommodate participant preferences. The FGDs lasted 90 minutes, on average.

**Table 1:**
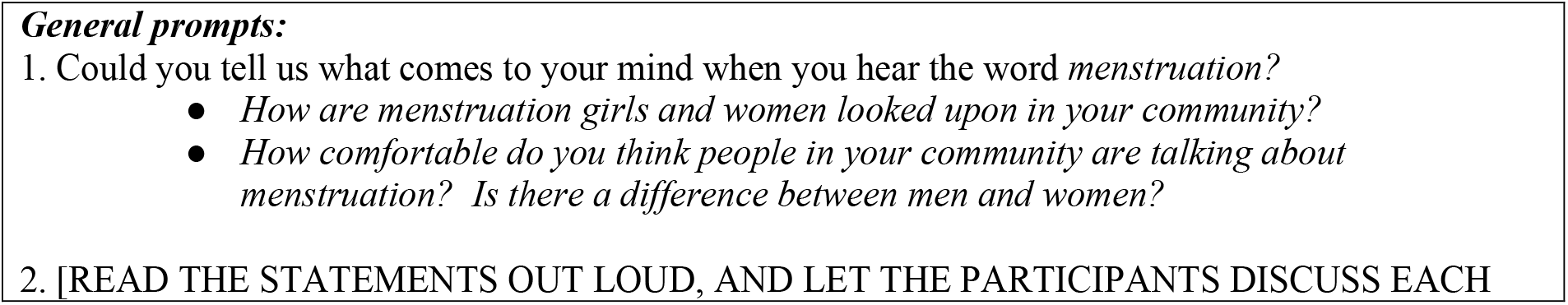
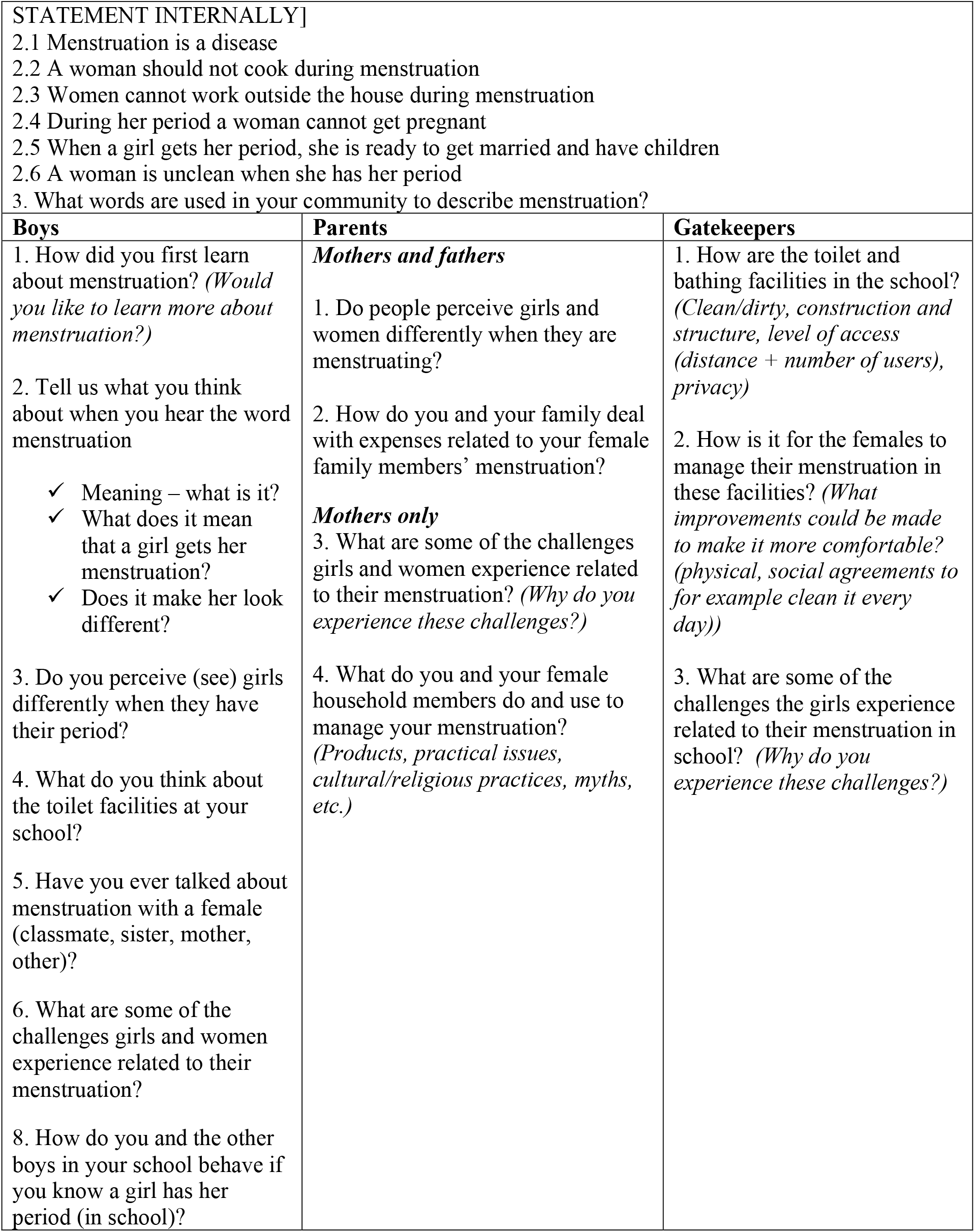
Focus Group Prompts.

### Data analysis

All FGDs were transcribed into text, with participant identities masked. Incoherent parts were repeatedly listened to and clarified for accuracy. Next, thematic analysis was conducted using MAXQDA 2024 software to generate codes, themes, and subthemes (27). Finally, specific examples were chosen for each theme. This study gathered perspectives from diverse participants (boys, parents, and various gatekeepers) to achieve “source triangulation” and ensure broad perspectives were captured on the topic of interest (28).

### Patient and Public Involvement

Patients and the public were not involved in any way in this research.

### Ethical considerations

Participation in the study was voluntary, and anonymity and the right to withdraw were honored. In addition, we collaborated with school officials to obtain formal (written) informed consent from parents for their children to participate, with written assent provided by the boys. Parents, teachers, and gatekeepers provided written informed consent before each FGD. There were no direct or reputational risks for individuals who participated in the research study, and participants’ confidentiality was strictly protected.

The Institutional Review Boards at UHAS (Study ID: UHAS-REC A.6 l2l23-24) and Michigan State University (Study ID: MOD00007866) reviewed and approved this study.

## Results

### Participant Demographics

The study involved 127 participants, categorized into three groups: adolescent boys, parents, and gatekeepers (see Table 2). The largest group consisted of 60 Muslim or Christian boys with varying ethnic backgrounds. The second group consisted of 48 parents, predominantly Ewe and Kotokoli, with varying educational backgrounds. The final group of 19 gatekeepers was mainly Christian and Ewe ethnic, with 74% working as teachers and others in education-related roles.

**Table 2:**
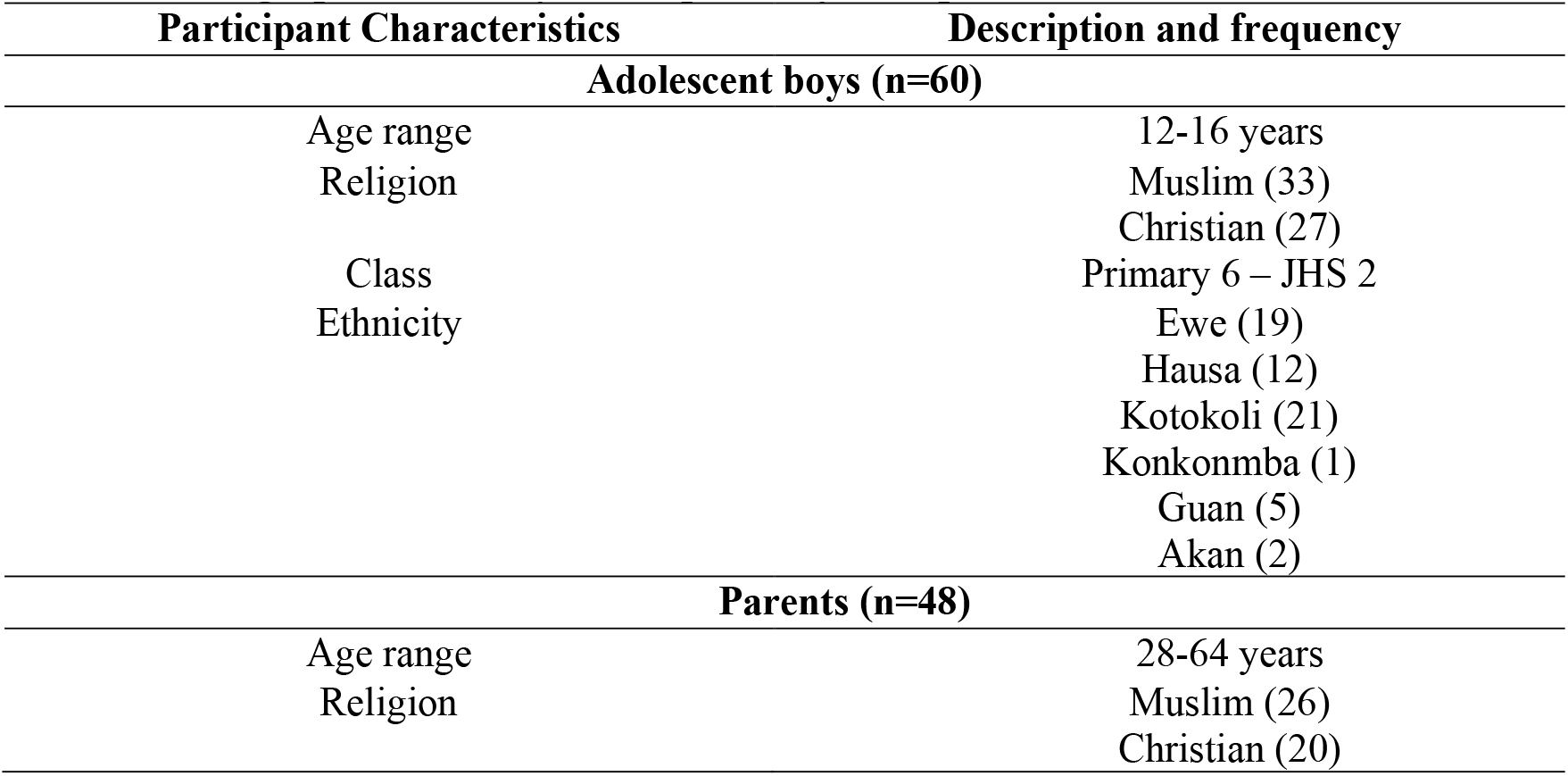
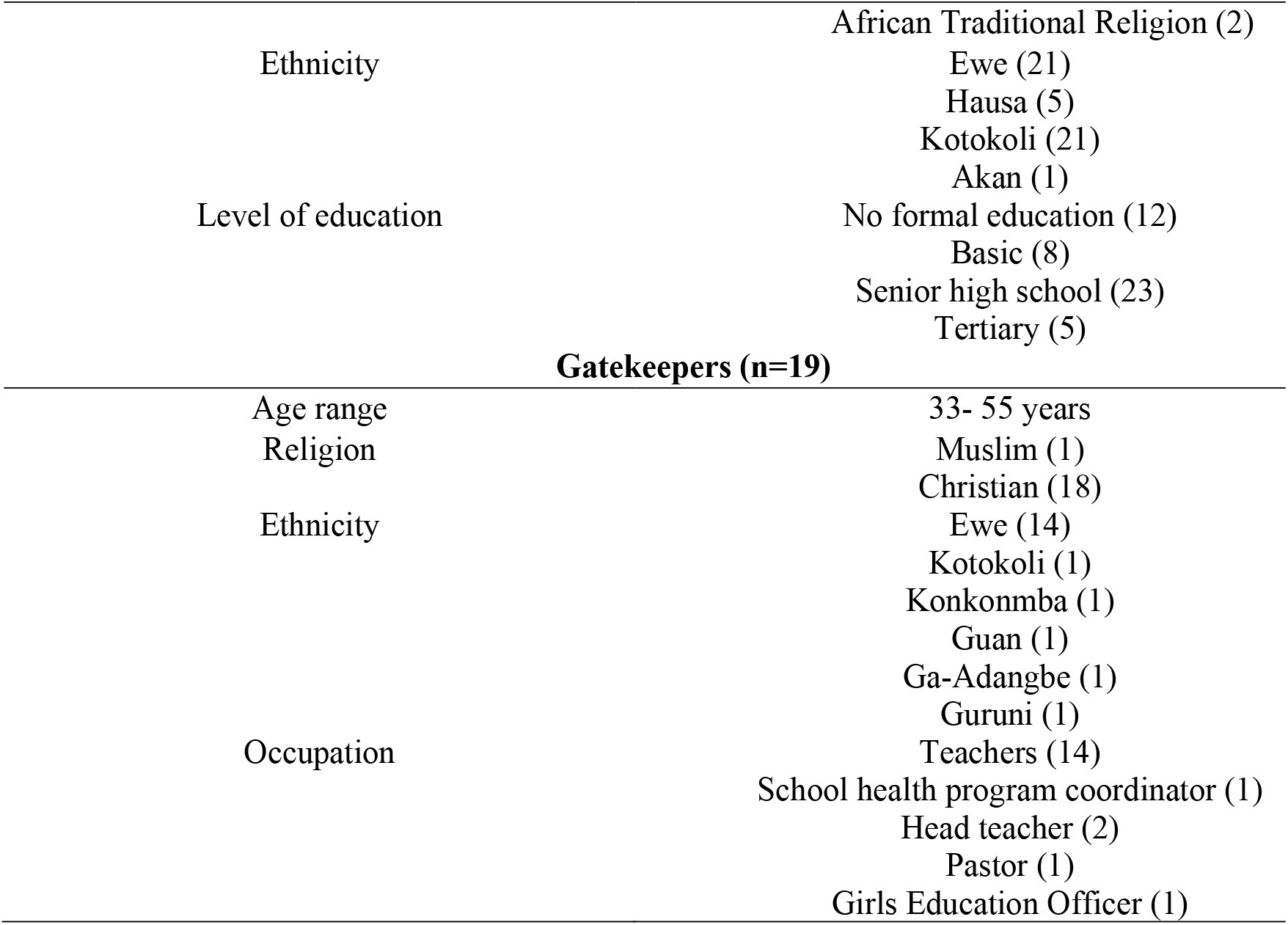
Demographics of Study Participants, by Group.

### Thematic results

The analysis generated major and minor themes, uniquely presented based on the three categories of participants. These themes are presented in Table 3 below.

**Table 3:** Themes Generated from the Interviews.

| Major themes | Minor themes |  |  |
| --- | --- | --- | --- |
|  | Parents | Schoolboys | Stakeholders |
| **Menstrual health management** | Menstrual pain management | Menstruation is not a disease | Management of menstrual cramps |
|  | Cost of menstrual products | Meaning of menstruation | Handling menstruation in school |
|  | Pain during menstruation | Perceived differences in girls during menstruation | Menstrual health recommendations |
|  |  | Pregnancy during menstruation | Prioritization of menstrual health expenses |
|  |  | First heard about menstruation |  |
| **Social and cultural perceptions** | Cultural perceptions towards menstruation | Cultural practices to manage menstruation | Age of menarche |
|  | Menstrual taboos and restrictions | Taboos associated with menstruation | Cultural relations and practices |
|  | Stigmatization and isolation | Home discussions about menstruation | Community and religious attitudes |
|  | Menstruating women are unclean | Boys' reactions to menstruating girls | Perceptions of menstruating women and girls |
| **Menstrual hygiene practices** | Bathing and sanitary practices | Materials used for menstrual management | Types of menstrual products |
|  | Use of disposable sanitary pad | Hygiene practices during menstruation | Keeping clean and WASH facilities |
|  | Use of alternative sanitary products | Toilet facilities | Rules about menstrual blood |
| **Education and awareness** | Comfort in discussing menstruation | Talking to girls about menstruation | Sharing of menstrual experiences |
|  | Shyness about menstruation | Comfortability in discussing menstruation | Sources of menstrual knowledge |
|  | Lack of awareness and taboos | Desire to learn more about menstruation | Need for more knowledge |
|  | Understanding of menstruation |  | Comfortability in discussing menstruation |

### Perspectives of Adolescent Boys

#### Perceptions and understanding of menstruation

Boys generally understand menstruation as a natural process for childbirth, not a disease. They perceive girls as sad, weak, and socially withdrawn during menstruation. Knowledge varies, with some learning from health workers, family, or school.

> “*Without it, women cannot give birth, is not a disease, is not a bad thing*.”
>
> “*Because it is the reason a girl can get pregnant*”.
>
> “*She can get pregnant during menstruating when she sleeps with someone*.”
>
> “*I first got to know about menstruation when some health worker came to our school to address it*.”

#### Social and cultural perceptions

Boys are aware of cultural practices related to managing menstruation, including traditional remedies like cooking unique leaves to reduce menstrual pain. They also acknowledge taboos that restrict menstruating girls from activities such as praying, cooking, fetching water, or participating in religious rituals. The quotes below summarize their responses.

> “*They are some leaves they cook for the girl to drink and sleep to avoid the pain*.”
>
> “*In my religion, women are not supposed to be praying when menstruating because she can’t play with the blood; they can’t go out*.”
>
> “*They are not allowed to cook, they can’t go out, can’t take part in sacrificial things, can’t work in the house, they cannot read the Quran, can’t fetch water from the river, men can’t sleep with you because they feel the Juju will work on you when you do that*.”

Discussions about menstruation at home are often minimal or conducted in secrecy, with boys learning from mothers or sisters. Their reactions to menstruating girls vary, ranging from teasing or avoidance to offering help. They show diverse attitudes based on their upbringing and personal views. The following quotes provide a summary of their responses.

> “*We don’t talk about menstruation at home because, when you ask a question, they don’t mind you; my sister and mother talk about it every day*”.
>
> “*I will be like, don’t you know you are menstruating? Then I will give you your privacy. When my friend sees they will start laughing and shouting eii eii hmm she is menstruating*”.
>
> “*Sometimes I want to help them, but I don’t know how to*”.

#### Menstrual hygiene practices

Regarding menstrual hygiene practices, boys know the materials used for menstrual management, including pads, rags, menstrual cups, and toilet rolls. They understand the importance of maintaining hygiene during menstruation, which includes regular bathing, washing clothes and cleaning any blood. The following quotes summarize their expressions.

> “*They use pad, toilet roll, pad, bags, wear pads*”.
>
> “*They wear pads, menstrual cups, hot water, medicine*.”
>
> “*Washing their clothes, washing pants, bath regularly, cleaning the blood*”.

However, most of the boys observe that toilet facilities in schools are often inadequate, unclean, and lack privacy, making it challenging for girls to manage menstruation discreetly. This recognition highlights the need for better infrastructure and resources to support school menstrual hygiene management. The quotes below reflect their responses.

> “*Very bad and dirty, you see pad and blood there Dirty*.”
>
> “*The teachers own [toilet facility] is good, the student own is not good. Sometimes you see blood on it. Teachers have water closets, but the students don’t. Kids defecate on the toilet every day. Teachers have dustbins, but students don’t have. Sometimes you see pads on the ground*”.

#### Education and Awareness

Most of the boys have mixed feelings about discussing menstruation with girls—some feel comfortable, while others feel shy or avoid the topic altogether. They are more at ease discussing menstruation with mothers, sisters, or close female friends. Despite this discomfort, many boys express a desire to learn more about menstruation, primarily to support their sisters or future daughters. This reflects a growing awareness among boys that understanding menstruation is essential for offering support and breaking down taboos, indicating a shift towards more open and informed perspectives. Below is how they expressed themselves.

> “*I talk to my sister about that, and it is normal; my friends feel ‘disgusted’ when I talk about menstruation to them, so I don’t*”.
>
> “*I feel shy because the boys say I’m not a girl*.”
>
> “*I want learn more because when I grow and born a child and when the child is a girl I can teach her. I have younger sister when she start menstruation I will teach her how to do anything…*”

### Perspectives of parents

#### Menstrual health management

This theme reveals how parents manage menstrual health. Mothers describe various ways of managing menstrual cramps, ranging from taking medication to resting, which indicates variability in pain management practices. However, some parents mentioned that they do not experience menstrual pain. These quotes below reveals what the parents said.

> “*I close my eyes in pain and try to sleep, and the whole house knows the rules, so no one should come and disturb me, and when I’m in the hall, and the pain is severe, and you’re watching television, you need to put it off because I get angry quickly*.”
>
> “*For me, I don’t have pain, but my daughters do, so I get them medications to stop the pain*.”
>
> “*The pain we go through and our daughters, we need medications to stop the pain*”.

The cost of menstrual pads is a significant challenge for some families, leading them to adopt alternative methods, such as using reusable cloths to manage menstruation at home. Others said they could afford the pads. The following quotes summarize their responses.

> “*So, my daughter and I bought it, and we struggled a lot because of the cost*.”
>
> “*For my side, when my kids are at home, I let them use the reusable ones since the pads are expensive, and I buy them the normal pads when they go to school*.”
>
> “*We buy it down, but because it can be finished anytime, I ask her to use either the clothes or the reusable ones*.”
>
> “*So, buying pads isn’t a problem for me*.”
>
> “*It disturbs me a lot because it is expensive, so at times, I buy it and put it down for her*.”

#### Cultural and social perceptions

The data reveals a shift in perceptions and practices surrounding menstruation across different cultural and religious groups. Traditional rituals and taboos, once rooted in Indigenous spiritual practices, have diminished significantly, influenced by modernization and the spread of religious practices that discourage Vudu and juju. However, certain restrictions persist, particularly those tied to spiritual practices, such as visiting places of worship while menstruating, and cultural norms, such as prohibitions on cooking or accessing specific spaces. This demonstrates the erosion of older traditions and the enduring nature of some beliefs. The quotes below summarize parents’ views on the topic.

> “*in the old days, our parents do Vudu and juju, so they don’t like it at all to see women menstruating come close to them krona. But now, because of Christianity, these things are over*”.
>
> “*So those days we do rituals for those menstruating but now we don’t do it and someone like me from krobo we do dipo rites but these things are no more*”.
>
> “*We the Muslims, when a woman gets her period, she doesn’t go the mosque to pray*”..
>
> “*It is still practiced that when you are menstruating you don’t cook for your husband or go to some certain places especially the chief palace*”.

In some communities, menstruating girls and women are still perceived as unclean or impure, reflecting persistent stigma. The following quotes summarizes their response.

> “*When women are menstruating in the society, we men see her some ways or we perceive them filthy, and we don’t want them to be among us and that’s how we see them*”.
>
> “*In my home, we use different buckets [to collect water] apart from men because we use the same buckets when we are in our periods, and we can’t be using the same bucket to fetch water*”.
>
> “*We see ladies that are menstruating to be dirty*.”

#### Menstrual hygiene practices

Parents emphasized personal hygiene practices during menstruation, such as regular bathing, changing pads, washing and drying undergarments, and maintaining cleanliness. They also mention some cultural or religious rituals, such as the specific bathing prayer for Muslim women. Limited financial resources sometimes force families to resort to alternative cleaning products like lemon to maintain hygiene during menstruation. The following quotes reflect the responses of parents.

> “*When it happens that way, I ask her to bathe, and when she is going out, I make sure she is wearing a sanitary pad, and when at home, she will wash her pants very well and dry it*”.
>
> “*When it happens, we take care of ourselves, bath regularly and change our pads*.”
>
> “*For we, the Muslims, when she is done with her menses, there’s a word she is supposed to say after bathing before she can pray*”.
>
> “*Asked her to use lemon in her armpit and to bath well since I don’t have money to buy Dettol always*.”

#### Menstrual education and awareness

Many mothers felt uncomfortable discussing their menstrual experiences, with some parents believing it is too personal or “dirty” to share beyond immediate family members like a husband or children. The discomfort is compounded by cultural norms and financial concerns, particularly when women struggle to afford pads. However, some parents do share menstrual information with their husbands to manage family planning and intimate relations. The negative stigma surrounding menstruation can also lead to poor hygiene practices, as described in one case of menstrual odor. These quotes summarize the parents’ responses.

> “*When people are having their menstruation, they are not comfortable speaking about it, and you can see they’re complaining about their abdomen, and some too are complaining about money to buy their pads*.”
>
> “*Menstruation is a shyness thing, so when people are menstruating the society, the way they will speak about it they and cannot also talk to people about it*.”
>
> “*As a woman, you can’t discuss issues of menstruation with your family. It is a matter of dirt, and no one needs to know about it; you must not tell anyone*”.

The majority of parents discussed a mix of biological, cultural, and spiritual understandings of menstruation. Many view menstruation as a natural, God-given process, where “bad blood” is discharged from the body to cleanse and prepare a woman’s womb. Menstruation marks a significant transition from childhood to womanhood, signaling a girl’s entrance into adolescence. Alongside the physical aspects, there is an emphasis on hygiene and teaching girls how to care for themselves during this time. Menstruation is also tied to family traditions, with many believing it has been a part of life for generations, passed down from ancestors. The following quotes reflect the responses of parents.

> “*Our mothers, women or siblings, women, every new month, bad blood is discharged from them, which is a natural thing set down by God*.”
>
> “*Is a time when a girl becomes an adolescent and they reach some certain age and when they reach that age blood comes from them*”.
>
> “*What I understand is gbototsitsi” that’s the bad blood that women have withing them comes out for them to be clean and prepared their wombs*”.
>
> “*If I hear the word Menstruation, what comes into my mind is that, at that time, what are the things I should do in other not to get pregnant during my period*”.

### Perspectives of gatekeepers

#### Menstrual health management

Most gatekeepers shared perspectives on managing cramps through medication or resting. In schools, poor hygiene facilities make handling menstruation difficult. Gatekeepers stressed the need for more education on menstrual health and the donation of menstrual products by government and NGOs. Financial concerns over buying menstrual products affect school attendance and the overall comfort of menstruating girls. The following are representative quotes from the gatekeepers:

> “*Yeah, we don’t have the toilet facility …, no changing room so they suffer when they are in their menses and then no toilet too, so they suffer*”.
>
> “*The men are not ready, but I think with more education, when we [teachers] educate them [males] on the good side of knowing these things, they [males] will be interested. For our men [male teachers], they can read on it and ask few questions from their mothers and partners*”.
>
> “*The government should reduce tax so that they will reduce the price of the sanitary pad. It is too expensive. They can’t buy it*”.

#### Cultural and social perceptions

This theme revealed cultural and social perceptions of menstruation among the stakeholders. Cultural practices, such as the Dipo rites and taboos, view menstruation as either a disease or impurity. Community and religious attitudes towards menstruation create discomfort, particularly among men, and vary across regions. Menstruating girls and women are sometimes stigmatized as unclean or impure, which negatively impacts their self-esteem and participation in community activities. The following quotes reflect the responses of participants.

> “*There is some this thing concerning this menstruation especially, among our traditional people, indigenous this thing. They have a culture whereby when you have your menstrual period for the first time, you have to go through some rites; the Dipo rites, the gboto …, and those rites, that is where they will get to know whether you have err, kept yourself till that time or, ahaa*”.
>
> “*Some churches, when you are menstruating, you are not allowed to enter and the shrine. Those who are wives to the custodian, the chief priest, their wives are not allowed to cook for them when they are menstruating*”.
>
> “*I think in the community, they leave it [menstruation] to women to talk about those issues. It’s not a topic that is been discussed outside. So, most a times, the women, when your child is menstruating, you must call the child and then talk to her. It’s not something that we talk about*”.

#### Menstrual hygiene practices

Most participants use a variety of menstrual products like pads, cloths, and rags due to limited resources. Maintaining hygiene is difficult, especially with inadequate water, sanitation and hygiene (WASH) facilities in schools. Participants also revealed that cultural rules such as disposal practices tied to rituals^2^ dictate how menstrual blood is handled, often adding to the challenges of menstrual management. The following quote summarizes the responses.

> “*My sister said we use red cloths those days because of fear but now because of pads they go anywhere*”.
>
> “*Yeah, because of the way and manner a lot of people want to get rich quickly, some use it for ritual purposes and err, go for some of these things. When you dispose your blood off, they see it then they use it for affectatious things*”.
>
> “*Truly, the toilet facilities for the kids, we don’t have it but sometimes, they go to the teachers’ [toilet facilities]. Theirs have spoilt and they couldn’t repair it for them so is a problem but when they go to the teachers’ place, hmm, they use it like that*.”

#### Education and awareness

This theme revealed that, education on menstruation is limited, with girls often refraining from sharing their experiences due to shyness or fear of teasing. Knowledge about menstruation comes from various sources, including parents, teachers, media, and menstrual hygiene clubs, but remains insufficient.

> “*It is not something our culture really makes it a topic to talk about at home but we have developed out of the era of um, looking down on menstrual this thing. People are becoming much aware of what it is. It is a natural phenomenon. Nothing can be done about it. So, it’s comfortable but we don’t make it a topic thing. We see it more of a woman thing so women are into more of their topic thing than we men. We don’t talk about it. We leave it to them to their own thing*”.
>
> “*Our society frowns on these kinds of things. So, it is not a topic we comfortably discuss in public. Even with, even*^***1***^ *as a lady, you can’t just say to your colleague ladies that today I’m menstruating. Are you getting it? It’s just maybe, a very close friend so it’s not a comfortable topic to discuss in our culture*”.

Stakeholders stated that, there is a recognized need for more comprehensive education, particularly for boys and men, to reduce the stigma around menstruation and encourage supportive interactions. Comfort levels in discussing menstruation vary widely, often influenced by cultural upbringing and the taboo nature of the subject, emphasizing the need for broader awareness and education initiatives. The quotes below summarizes their responses.

> “*More awareness should be created for stakeholders involved to support the girls to take these materials, get the materials and use because is natural something. Whether they like it or not, it will continue to come. So, how we should support them to get these items is over to the government and stakeholders*”
>
> “*I have all boys. All my children are boys. So, there was a time one of them came to ask me; how do women get pregnant? So, this is not just a straightforward question, is a broader thing so menstrual this thing has to come so I sat them down and teach them the menstrual process, the attitudes about them, sex, I had to teach them about it and it was a whole lot. They asked, they wanted to know and I couldn’t tell them anything so I taught them what it is and extra care also came in*”.

## Discussion

This study explored stakeholders’ perspectives on improving menstrual health and hygiene for school-going adolescent girls in Hohoe Municipality, Ghana. Key stakeholders, including adolescent boys, parents, and community gatekeepers, highlighted several critical issues: varying understanding of menstruation, persistent cultural taboos and restrictions, challenges in menstrual hygiene management, inadequate educational resources and awareness, and the need for improved facilities and support systems. Boys wanted to support their female peers, while parents and gatekeepers emphasized the importance of education and the need to address financial barriers to accessing menstrual products. The findings also highlighted the impact of cultural beliefs on menstrual practices and the role of comprehensive education in breaking down stigma and improving menstrual health experiences for adolescent girls.

The findings underscore the significant barriers to menstrual health and hygiene (MHH) in Hohoe’s schools, such as product inaccessibility, inadequate facilities, cultural stigmas, and limited education, consistent with reports from other areas in Ghana. (3,29). Participants noted insufficient WASH facilities and cultural stigma on girls’ attendance and participation in school findings reported elsewhere. (3,30)

Financial constraints compel girls to use unsafe alternatives, such as rags and tissue paper (31,32,33,34), and suggestions for tax reductions on menstrual products could alleviate this burden and echo discussions on their affordability. (32,35). However, further research is needed to assess the impact of such policy changes.

Participants emphasized the lack of adequate WASH facilities in schools, a challenge noted in other sub-Saharan African studies ((36–38). The absence of private, clean toilets and changing spaces hinders effective menstrual hygiene management, contributing to girls’ discomfort and absenteeism. These perspectives align with research suggesting that inadequate school infrastructure disproportionately affects menstruating girls (39).

Additionally, participants noted cultural and social taboos surrounding menstruation, many of which are consistent with earlier studies (4,40). Some respondents noted that girls who menstruate are often excluded from religious or communal activities, which resonates with findings from a study in Northern Ghana (3). Cultural beliefs that view menstruation as “impure” impact discussions around menstruation, affecting both girls and boys’ understanding, reinforcing gender-based misconceptions. Meanwhile, parents, particularly fathers, expressed discomfort discussing menstruation, which reflects a broader trend of parental reticence found in previous research (22).

Parents and gatekeepers highlight the need for more comprehensive menstrual health education. Participants indicated that current educational efforts are insufficient, which aligns with findings from Sommer et al. (39), who argue that both boys and girls would benefit from integrated menstrual health education in school curricula. Gatekeepers expressed a need for more resources and training for teachers to properly educate students, consistent with findings from UNESCO (41) suggesting that teachers often lack the necessary materials to educate students.

Engaging boys in menstrual health education is crucial to reduce stigma and promote more supportive interactions which can help dismantle stigmatizing attitudes (3,22). It was encouraging that boys mentioned they would like to “help girls but don’t know how”. Boys indicated a desire for guidance on assisting their peers, emphasizing the importance of education and mentorship.

### Strengths of the study

The study excels at stakeholder engagement by involving boys, parents, and gatekeepers (predominantly educators), resulting in a complete understanding of barriers to menstrual health management. Engaging boys is critical for challenging gender norms and creating a welcoming environment for girls. It is also contextually relevant, focusing on Hohoe’s unique cultural and socioeconomic environment. This local perspective enables the study to address the unique challenges that adolescents in the area face and identify key barriers, such as cultural stigmas and inadequate facilities, which can inform targeted interventions.

### Study limitations

The study’s emphasis on Hohoe limits its generalizability, as it may not apply to other parts of Ghana or elsewhere where cultural and demographic differences exist. There is also the possibility of selection bias, as participants who are more concerned with menstrual health issues may have participated more frequently, potentially skewing the results.

## Conclusion

This study examined barriers and strategies to improve menstrual health and hygiene (MHH) for school-going girls in Hohoe, Ghana, through the perspectives of key stakeholders. Cultural stigmas, inadequate facilities, and financial constraints emerged as major challenges, highlighting the need for a comprehensive approach involving boys, parents, and gatekeepers to break taboos and foster open dialogue. Recommendations include implementing school menstrual health education programs, engaging both boys and girls and involving parents to better support their daughters. Additionally, improving school WASH facilities with private toilets, changing rooms, and proper disposal systems is crucial. There is a need to reduce financial barriers through the reduction of taxes on sanitary products, subsidizing kits, or promoting reusable options like menstrual cups to enhance access and reduce absenteeism, and create a more supportive environment for girls.

## Data Availability

All relevant data are within the manuscript. Any further requests regarding the data used for this study can be made through the corresponding author.

## Acknowledgements

We thank the school health education program coordinators who assisted in the study.

## Declarations

### Corresponding Author

The Corresponding Author has the right to grant on behalf of all authors and does grant on behalf of all authors, an exclusive licence on a worldwide basis to the BMJ Publishing Group Ltd to permit this article (if accepted) to be published in BMJ editions and any other BMJPGL products and sublicences such use and exploit all subsidiary rights.

### Competing Interest Statement

All authors have completed the <u>Unified Competing Interest form</u> and declare: $5,000 was received from the Consortium of Universities for Global Health to support this work; no other relationships, payments or activities have influenced the submitted work.

### Transparency Declaration

The lead author (the manuscript’s guarantor) affirms that the manuscript is an honest, accurate, and transparent account of the study being reported; that no important aspects of the study have been omitted; and that any discrepancies from the study as planned have been explained.

### Funding Acknowledgement

This study was funded by a Consortium of Universities for Global Health Tom Hall Education Grant. Neither the authors nor their institutions at any time received payment or services from a third party for any aspect of the submitted work.

### Independence of Researchers from Funders

research findings and conclusions are solely based on the data and analysis, without influence or pressure from the Consortium of Universities for Global Health.

### Author Contributions

Conceptualization: S.G., C.C. and I.W.; Methodology: S.G., V.C.O. and I.W.; Formal analysis: I.W., P.K., S.O.M., and I.B; Writing-original draft: I.W., P.K., and S.O.M.; Writing-review and editing: D.E., C.C., S.G., I.W., P.K., I.B., S.O.M., and P.E.N.; Project supervision: S.G., and V.C.O.; Project administration: P.E.N., P.K., I.W, I.B., and S.O.M. All authors have read and agreed to the published version of the manuscript.

### Conflicts of Interest

The authors declare no conflicts of interest.

### Patient and public involvement

Patients and/or the public were not involved in the design, conduct, reporting, or dissemination plans of this research.

### Ethics Statement

The study was conducted in accordance with the Declaration of Helsinki and approved by the University of Health and Allied Sciences Research Ethics Committee (Study ID: UHAS-REC A.6 l2l23-24) and Michigan State University (Study ID: MOD00007866).

## Footnotes

1 Menstrual blood is believed to be powerful and being used for rituals which affect how women dispose of their menstrual absorbents.

## Notes

### Competing Interest Statement

The authors have declared no competing interest.

### Funding Statement

This study received $5,000 from a Tom Hall Education Grant, Consortium for Universities in Global Health.

### Author Declarations

The Human Research Protection Program of Michigan State University gave ethical approval for this work ((Study ID: MOD00007866). The University of Health and Allied Sciences Research Ethics Committee gave ethical approval for this work (Study ID: UHAS-REC A.6 l2l23-24).

